# Integrating mouthguard kinematics, finite element brain strain, and plasma biomarkers to explore brain injury thresholds in collision sport

**DOI:** 10.64898/2026.08.26.26360869

**Authors:** James W. Hickey, Emily Yik Kwan Chan, Lauren J. Evans, William T. O’Brien, Becca Xie, Spencer S.H. Roberts, Sarah E Butler, Joel Ernest, William Ji Quan Zhou, Karl A. Zimmerman, Gershon Spitz, Thomas D. Parker, Terence J. O’Brien, Sandy R. Shultz, David J. Sharp, Mazdak Ghajari, Stuart J. McDonald

## Abstract

**Purpose:** Identifying head impacts linked to brain injury in sport remains challenging. Instrumented mouthguards quantify head-impact kinematics, and finite element (FE) modelling can transform these data into brain strain estimates, which may better reflect injury risk than kinematics alone. Here, we examined associations between mouthguard-measured kinematics, FE-derived strain, and plasma brain injury biomarker GFAP following head impacts.

**Methods:** We analysed 41 video-verified impacts from male Australian football players, including 22 assessed for concussion (17 diagnosed) and 19 unassessed. Instrumented mouthguards recorded peak linear acceleration (PLA), peak rotational acceleration, and peak rotational velocity (PRV). Brain strain was estimated using the Imperial College FE brain model, and plasma GFAP was quantified using Simoa. Biomechanical-GFAP associations were examined using Spearman correlations and segmented regression.

**Results:** For impacts overall, plasma GFAP was moderately correlated with PLA (ρ=0.46, 95% CI: 0.20–0.66), PRV (ρ=0.53, 95% CI: 0.20–0.78), and strain (ρ=0.60, 95% CI: 0.32–0.80). Associations were stronger within concussion cases for strain (ρ=0.86, 95% CI: 0.58–0.97) and PRV (ρ=0.64, 95% CI: 0.15–0.93). Piecewise regression identified strain levels above which strain-GFAP relationships steepened across the whole-brain and brainstem. In concussion cases, supra-threshold brainstem strain was associated with greater symptoms.

**Conclusion:** Finite element brain strain may better predict brain injury risk following a sport-related head impact than peak acceleration metrics. Stronger associations with plasma GFAP, particularly among concussion cases, and evidence of a biomechanical threshold, support the use of biomarker-informed strain measures in future risk modelling and the development of brain injury screening thresholds.

## Introduction

Identifying head acceleration events that carry a high risk of traumatic brain injury in sport remains a major challenge. These injuries arise when external forces to the head or body generate translational and rotational motion of the head, leading to brain tissue strain and microstructural disruption^1^. Concussion is the most common clinical manifestation of brain injury in sport^2^. Accurately detecting sport-related concussion, however, is difficult as neurological signs may be missed by sideline staff and symptoms may be under-reported and are inherently non-specific to brain injury^3–5^. Moreover, even when accurately identified, these clinical indicators do not reliably reflect the presence or severity of underlying brain injury^6,7^. This underscores the need for more objective approaches to predict and detect brain injury risk in sport, to support timely identification and management and reduce the risk of prolonged recovery or further injury.

Instrumented mouthguards (iMGs) now enable high-resolution recordings of head impact kinematics - including peak linear acceleration (PLA), peak rotational acceleration (PRA) and peak rotational velocity (PRV) - demonstrating strong accuracy and reliability in both laboratory and field settings^8–11^. World Rugby have recently implemented PLA and PRA thresholds to trigger a head injury assessment (HIA) in elite rugby union^12^. However, selecting an arbitrary universal threshold is challenging as it often fails to correspond with concussion diagnosis and may not reflect the presence of underlying brain injury. Whilst informative, these kinematic metrics describe only the external motion of the head, and their application may be limited as they do not account for the complex interplay of biomechanical forces and internal responses that occur within the brain following an impact.

Finite element (FE) models represent a promising solution to this limitation. By simulating the mechanical response of brain tissue to head impact kinematics recorded by an iMG, FE models can generate anatomically specific estimates of tissue strain^13–20^, which is thought to better reflect injury-relevant loading. Among available FE models, the Imperial College London model^18^ offers one of the highest levels of anatomical resolution and fidelity. It includes over one million hexahedral brain voxels, enabling highly precise spatial characterisation of internal brain deformation and voxel-level computation of maximum principal strain, which is a measure of maximal local brain deformation. This model has shown that brain strain is concentrated in the depths of sulci in sports collisions, aligning with the location of pathology seen in chronic traumatic encephalopathy^18^. It has also been used to predict the location of microbleeds attributed to a rugby tackle^21^, and that loss of consciousness in sport-related concussion is associated with high strain rates within the brainstem^19^.

Despite the promise of kinematics and FE-derived brain strain metrics, most studies evaluating their predictive value have relied on the binary classification of concussion versus no concussion as the primary outcome^13,14^. This approach carries several limitations. First, clinical diagnosis is inherently subjective, influenced by factors such as observer variability, self-report biases, and differing interpretations of subjective diagnostic thresholds^22,23^. Second, the presence or severity of symptoms does not necessarily reflect the extent of underlying tissue disruption, as symptom expression can be influenced by multiple pre-injury factors^24^ and may instead reflect cervical injury, peripheral vestibular disturbance or physiological stressors^25,26^. Third, the use of a binary outcome constrains the ability to explore dose-response relationships between biomechanical loading and injury severity, limiting insights into the continuum of brain injury risk.

An emerging approach for quantifying the presence and severity of traumatic brain injury involves the use of blood-based biomarkers. Glial fibrillary acidic protein (GFAP) and neurofilament light (NfL) are structural proteins that are released into peripheral circulation following astroglial injury/reactivity and axonal injury, respectively. GFAP is released from astrocytes, with levels typically peaking around 16-24 hours after concussion^27^, whereas NfL rises more gradually and has a longer half-life, reaching peak concentrations approximately 1-3 weeks after injury^6,27,28^. While we have recently reported evidence that iMG-derived PLA and PRA were associated with post-match GFAP and NfL elevations^29^, this analysis focused exclusively on non-concussive impacts, limiting the range of impact magnitudes and likelihood of injury studied. Moreover, no studies have systematically evaluated how kinematic or FE-predicted brain strain measures relate to biomarker evidence of brain injury following concussion, leaving a critical gap in understanding which biomechanical parameters best reflect underlying tissue-level injury in this context.

Accordingly, to our knowledge, we present the first biomechanics-biomarker study including diagnosed sport-related concussion, linking video-verified impacts and iMG kinematics to plasma GFAP and NfL. We extend this by incorporating FE-predicted brain strain to identify which measures best reflect brain tissue injury. The overall aim was to characterise the relationships between iMG-derived kinematic measures of PLA, PRA and PRV, and FE-estimated brain strain, with plasma concentrations of GFAP and NfL across a broad range of impact severities. To achieve this, we sampled impacts in Australian football spanning i) non-concussive events, ii) incidents that prompted a head injury assessment but were medically cleared, and iii) verified concussive impacts, capturing a wide range of biomechanical exposures. We hypothesised that brain strain would demonstrate stronger associations with biomarker levels than kinematic variables, reflecting the FE model’s more accurate representation of brain tissue deformation. We further hypothesised that these associations would be strongest among concussed athletes, as such impacts are more likely to exceed a tissue-disruption threshold sufficient to elicit a measurable biomarker response. Across impacts overall, we hypothesised that biomechanics-biomarker relationships may be non-linear, consistent with a biomechanical tipping point above which biomarker responses increase with mechanical loading. Finally, we examined whether identified breakpoints related to symptom burden among concussed athletes.

## Methods

### Standard protocol approvals, registrations, and patient consents

This prospective cohort study was conducted using data from multiple projects approved by the Monash University Research Ethics Committee (Project IDs: 27684, 36297, 36556). Written informed consent was obtained from all participants prior to participation.

### Recruitment and kinematic data processing

In total, 204 male amateur Australian football players from 11 teams underwent dental scanning to create custom-fit iMGs (HIT-IQ Pty Ltd, Melbourne, Australia) for use across the 2022-2024 seasons (***Figure 1***). In 2022, players (*n* = 21) wore Nexus Gen II iMGs, whereas 2023-2024 players (*n* = 183) wore the Gen III model. Both contained the same triaxial accelerometer (Analog Devices ADXL372; 3,200 Hz, ±200 g, 12-bit) and gyroscope (Bosch BMI270; 800 Hz, ±2,000°/s, 16-bit). Trigger thresholds differed slightly between models (Gen II: 13 g; Gen III: 8 g); however, all captures underwent identical post-processing, and this difference was deemed negligible given the study focus on higher-magnitude impacts. Kinematic data were recorded from 20 ms before initial trigger to 80 ms after the final trigger. The retrigger function enabled variable impact durations to be captured^30^.

**Figure 1.**
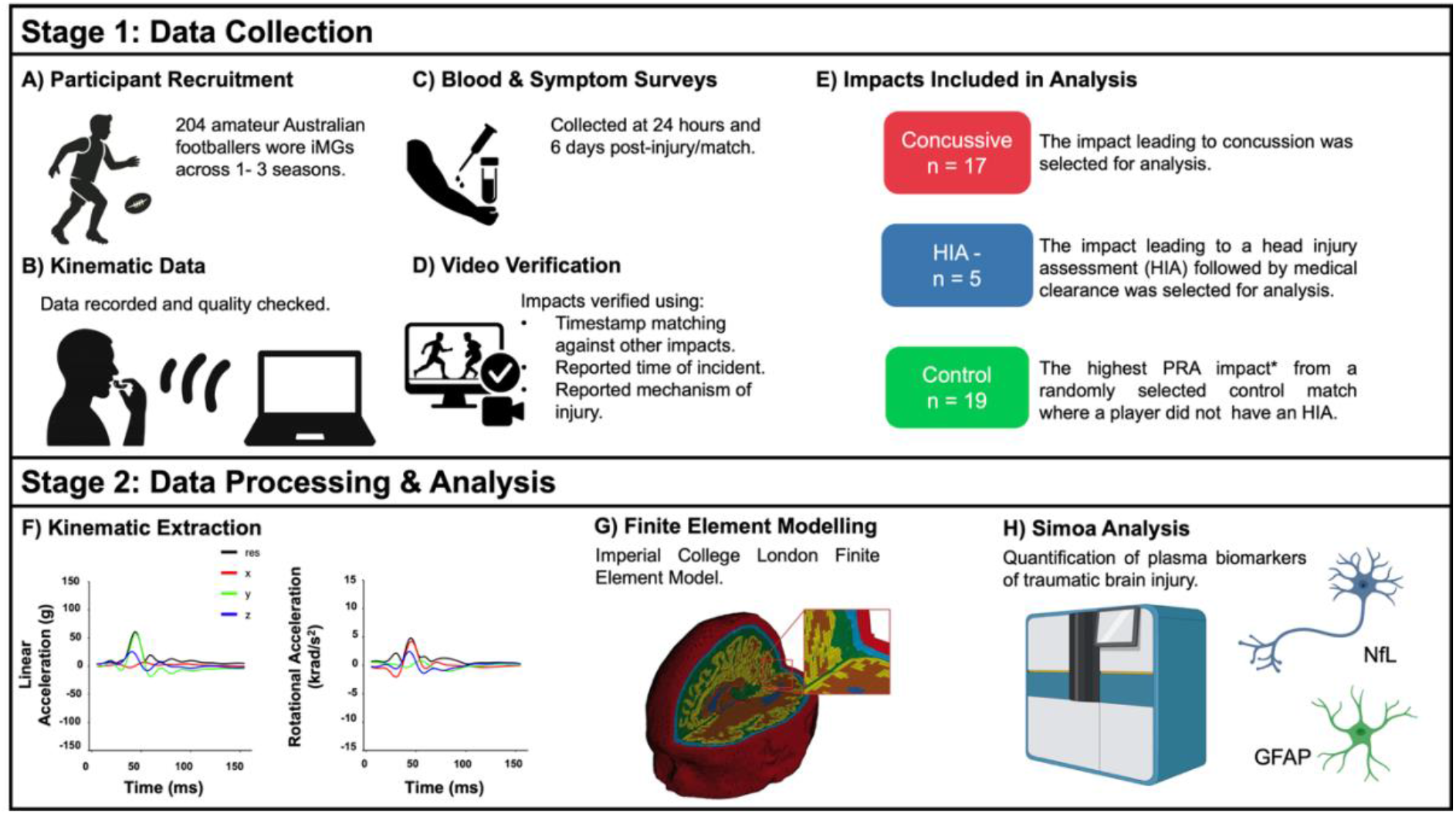
Data Collection and Processing Pipeline: (**A**) Two hundred and four male amateur Australian football players wore instrumented mouthguards (iMGs) during matches (2022– 2024). (**B**) Kinematic data were uploaded and extracted from HIT-IQ’s Nexus Portal. (**C**) Venous blood and symptom surveys were collected at 24 hour and 6 days post-match/injury. (**D**) iMG-recorded impacts were verified by two raters (**E**) Final participant sample included concussion (n = 17), medically cleared head injury assessment (HIA-) (n = 5), and control (n = 19). (**F**) Kinematic data were processed and verified using the same post-impact processing pipeline. (**G**) Finite element modelling was conducted using Imperial College London’s model; tissue segmentation includes skin (red), skull (light blue), cerebrospinal fluid (green), grey-matter (yellow), white-matter (brown), and ventricles (dark blue). (**H**) Plasma glial fibrillary acidic protein (GFAP) and neurofilament light (NfL) concentrations were quantified using a Simoa HD-X Analyser. *Alternative control impact selection methods were also explored and presented in Supplementary *Table 1*.

Participating teams provided post-match reports identifying players with a medically diagnosed concussion or those cleared following a sideline HIA (group termed HIA-) using the Sport Concussion Assessment Tool (SCAT). A similar number of control players who completed a match without concussion or HIA were recruited from a parallel study of non-concussive impacts and blood biomarkers within the same cohort and underwent identical testing. Controls were randomly selected to match concussion cases by age and season timing, with investigators blinded to impact and biomarker data. The impact leading to the concussion or HIA was verified using match footage and medical reports. Corresponding iMG data were accessed via HIT-IQ’s Nexus portal and verified against time-matched footage, reviewed by two raters.

To isolate the biomechanical effects of discrete head impacts, one impact per player match was included in the primary analysis. For concussion and HIA-cases, the impact corresponding to the documented event was analysed. For controls, the highest-PRA impact from the selected match was chosen based on an a priori hypothesis that PRA would best reflect brain strain and biomarker response (alternative selection methods reported in ***Supplementary Table 1***). Focusing on a single impact per player provided a practical framework to examine biomechanical drivers of biomarker changes and supports the development of individualised, event-level risk prediction using iMG data. Supporting this approach, our previous work on non-concussive impacts demonstrated that the maximum single-impact PLA in Australian football was associated with 24 hour GFAP levels^29^. Australian football may be suited to such analysis, as athletes typically sustain fewer impacts per match than in some other collision sports^31^, reducing potential confounding from repetitive exposures. Nonetheless, cumulative kinematic and brain strain metrics were also explored.

### Finite element modelling

Head impact kinematics were simulated using the Imperial College FE brain model, which incorporates detailed representations of cerebrospinal fluid, pia mater, grey-and white-matter, brainstem, ventricles, falx and tentorium. The model comprises approximately one million hexahedral and 250,000 quadrilateral elements derived from high-resolution magnetic resonance images of a healthy young adult male^18^. Measured linear and rotational accelerations were applied to the rigid skull (referenced to the centre of gravity), and all simulations were run using an explicit FE solver (LS-DYNA R9.1.0, LSTC, CA, USA). For each simulation, the maximum principal Green-Lagrange strain tensor (strain) and its time derivative (strain rate) were calculated for every element. Strain is a measure of an element’s maximum deformation and strain rate a measure of its maximum rate of deformation. Strain data were output in NIfTI (Neuroimaging Informatics Technology Initiative) format and spatially aligned to MNI152 (Montreal Neurological Institute) standard space using affine registration to enable anatomical consistency. The 90th percentile strain for the whole-brain, white-matter, grey-matter, and brainstem were extracted using binary masks generated from the MNI152 template using FAST segmentation^32,33^.

Group-average strain maps were obtained by aggregating voxel-wise strain across all impacts in each group and dividing by the total number of impacts. Rather than summing scalar strain which may overlook regional variations, this method preserves spatial information and regional patterns across the brain. Voxel-wise non-parametric permutation testing with FMRIB’s Software Library (FSL) randomise^34^ was used to assess group differences for both strain and strain rate, with distribution images concatenated across participants and analysed by a general linear model encoding group indicators. For each contrast, 10,000 permutations were run with image-wise maximal statistics to control the family-wise error (FWE) rate. Inference used Threshold-Free Cluster Enhancement (TFCE), and results are reported as TFCE-based, FWE-corrected *P*-value maps (*P* < 0.05).

For cumulative exposure, peak kinematic values were summed across impacts; cumulative strain was derived by summing voxel-wise strain maps across impacts to form a cumulative strain distribution, from which the 90th percentile was extracted. All image processing used FSL version 6.0.7.

### Blood collection and biomarker analysis

Biomarkers were sampled at timepoints aligned with their expected peak concentrations - GFAP at 24 hours and NfL at six days post-injury^6,27,28^. Six days was chosen to enable collection prior to a subsequent match. Venous blood was collected into K2EDTA tubes and stored at 4°C before centrifugation at 1100 *g* for 10 minutes. Plasma aliquots were stored at-80°C until batch analysis at the end of each season. Plasma GFAP and NfL were quantified using Simoa Neurology 2-Plex B Advantage kits on a Simoa HD-X Analyzer (Quanterix Corp., Billerica MA, USA). Samples were tested in duplicate with mean coefficients of variation of 8.5% for GFAP and 7.8% NfL.

### Statistical analysis

Analyses were performed using R (version 4.4.2). Data normality was assessed using visual inspection and Shapiro-Wilk tests, with parametric or non-parametric tests applied appropriately. Biomarker and biomechanical measures are reported as median and interquartile range (IQR). Group differences were tested using Kruskal-Wallis and Mann-Whitney U tests, with Holm-correction. Associations between biomechanical metrics and biomarkers were examined using Spearman’s correlations due to the positive skew of biomarker data. Bootstrapping (1,000 iterations) was used to generate 95% confidence intervals. Steiger’s Z test compared the strength of correlations. Non-linear associations were visualised using generalised additive models. Linear regression models adjusting for age, body mass index (BMI) and time-since-injury/match assessed associations between biomechanical measures and natural log-transformed biomarkers.

Breakpoint detection was performed using single-knot piecewise linear models with natural log-transformed biomarkers. Breakpoints were identified via grid search across the 10^th^-90^th^ percentiles (≥ 10 observations per segment), selecting the model with the lowest Bayesian information criterion and benchmarking against single-slope models using Akaike information criterion and 5-fold cross-validated root mean squared error. The *segmented* package was used to validate breakpoint estimates^35^. Changes in slope were tested using wild bootstrapping (10,000 Rademacher resamples). Model diagnostics were assessed using the *performance* package. Within concussion cases, Mann-Whitney U tests compared symptom severity above and below each breakpoint.

## Data availability

De-identified data and supporting code used for statistical analysis may be made available upon reasonable request.

## Results

### Participant characteristics, biomarkers, and biomechanical measures by sampling stratum

Of the 204 consenting players, 30 sustained a diagnosed concussion, and 5 underwent an HIA followed by medical clearance (HIA-). iMG data were available for 23 concussion and 5 HIA-events (***Supplementary Figure 1***). Match footage was available for video verification of 19 concussion and 5 HIA-cases; players without footage were excluded. Of these, 17 concussions and 5 HIA-cases had blood collected at 24 hours and six days post-injury. Final groups for analysis included concussion (*n* = 17), HIA-(*n* = 5), and control (*n* = 19) impacts (***Figure 1***).

Clinical strata (concussion, HIA-, control) were used for recruitment and data sourcing to sample impacts spanning routine match play through medically assessed incidents and clinically confirmed concussion. Descriptive summaries by stratum are provided in ***Supplementary Table 2*** and ***Supplementary Figure 2***. Briefly, median 24-h GFAP was higher following clinically assessed impacts, whereas 6-day NfL did not differ across strata. Kinematic measures and FE-derived strain were higher in concussion and HIA-impacts than controls. A progressive increase in the magnitude and spatial extent of strain was observed across groups, from control to HIA-and concussion impacts (***Figure 2***). Voxel-wise permutation testing further demonstrated higher strain and strain-rates in concussive impacts compared with non-concussive impacts (controls and HIA-) (***Supplementary Figure 3***). Strain differences were widespread across cortical and subcortical regions, whereas strain rate differences showed a more central distribution.

**Figure 2.**
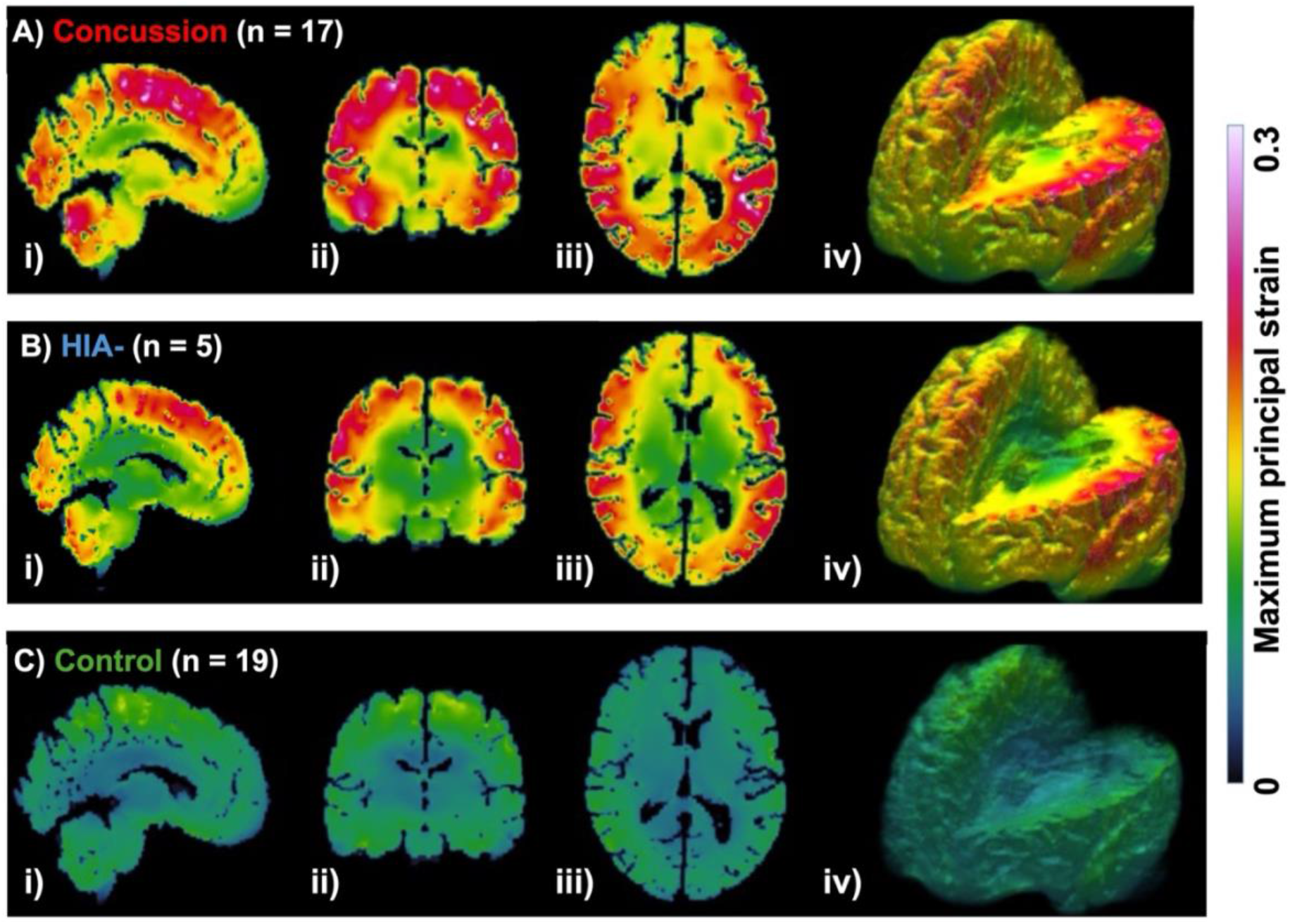
Spatial distribution by recruitment strata: Maximum principal Green-Lagrange strain (strain) are shown for impacts resulting in concussion (**A**; red; n = 17), medically cleared head injury assessment (HIA-; **B**; blue; n = 5), and control impacts (**C**; green; n = 19). Strain is displayed on a continuous scale from 0 (dark blue) to 0.3 (bright pink) for sagittal (**i**), coronal (**ii**), axial (**iii**) and 3D cortical surfaces (**iv**).

**Figure 3.**
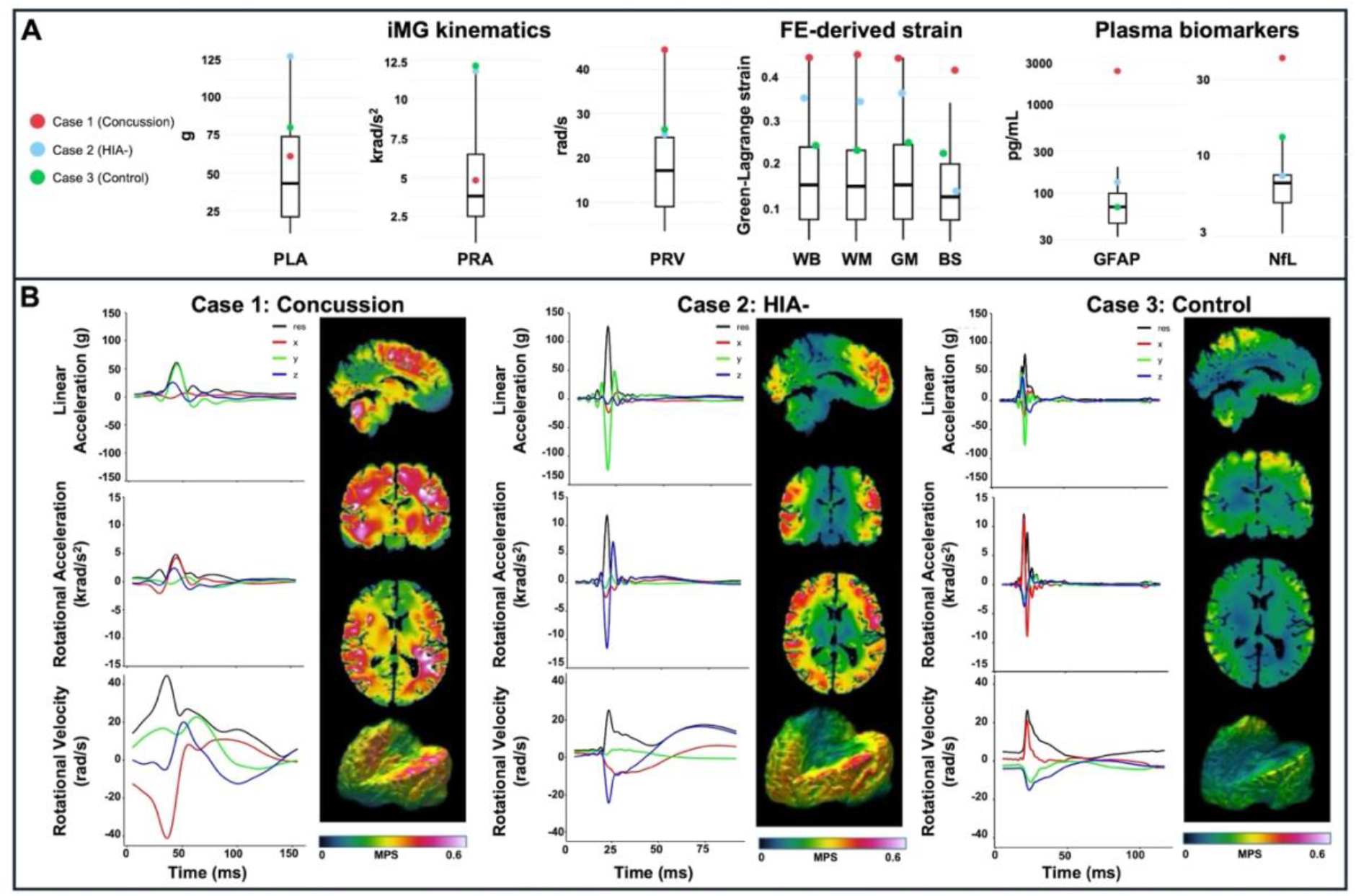
Case-level comparison of head impact kinematics, brain strain, and blood biomarkers. (**A**) Distributions of peak linear acceleration (PLA), peak rotational acceleration (PRA), peak rotational velocity (PRV) and finite element (FE)-derived maximum principal Green-Lagrange strain in the whole-brain (WB), white-matter (WM), grey-matter (GM), and brainstem (BS), and plasma glial fibrillary acidic protein (GFAP) and neurofilament light (NfL). Box plots show medians and interquartile ranges; whiskers extending to 1.5x the interquartile range for all participants; points highlight three representative cases with the highest strain within each group: concussion (Case 1; red), medically cleared head injury assessment (HIA-; Case 2; blue), and control (Case 3; green). (**B**) Time-series traces of linear acceleration, rotational acceleration, and rotational velocity components (x, y, z) recorded by instrumented mouthguard (iMG) for each case, shown alongside corresponding brain strain maps.

### Biomechanical associations with plasma GFAP

Moderate correlations were observed between GFAP levels and several biomechanical measures for impacts overall (***Table 1;*** Bonferroni-adjusted α = 0.006), including PLA (ρ = 0.46, 95% CI: 0.20–0.66, *P* = 0.003), PRV (ρ = 0.53, 95% CI: 0.20–0.78, *P* < 0.001) and whole-brain strain (ρ = 0.60, 95% CI: 0.32–0.80, *P* < 0.001) (***Figure 4***). Similar associations were found for white-matter, grey-matter and brainstem strain, as well as strain rate (***Table 1***).

**Figure 4.**
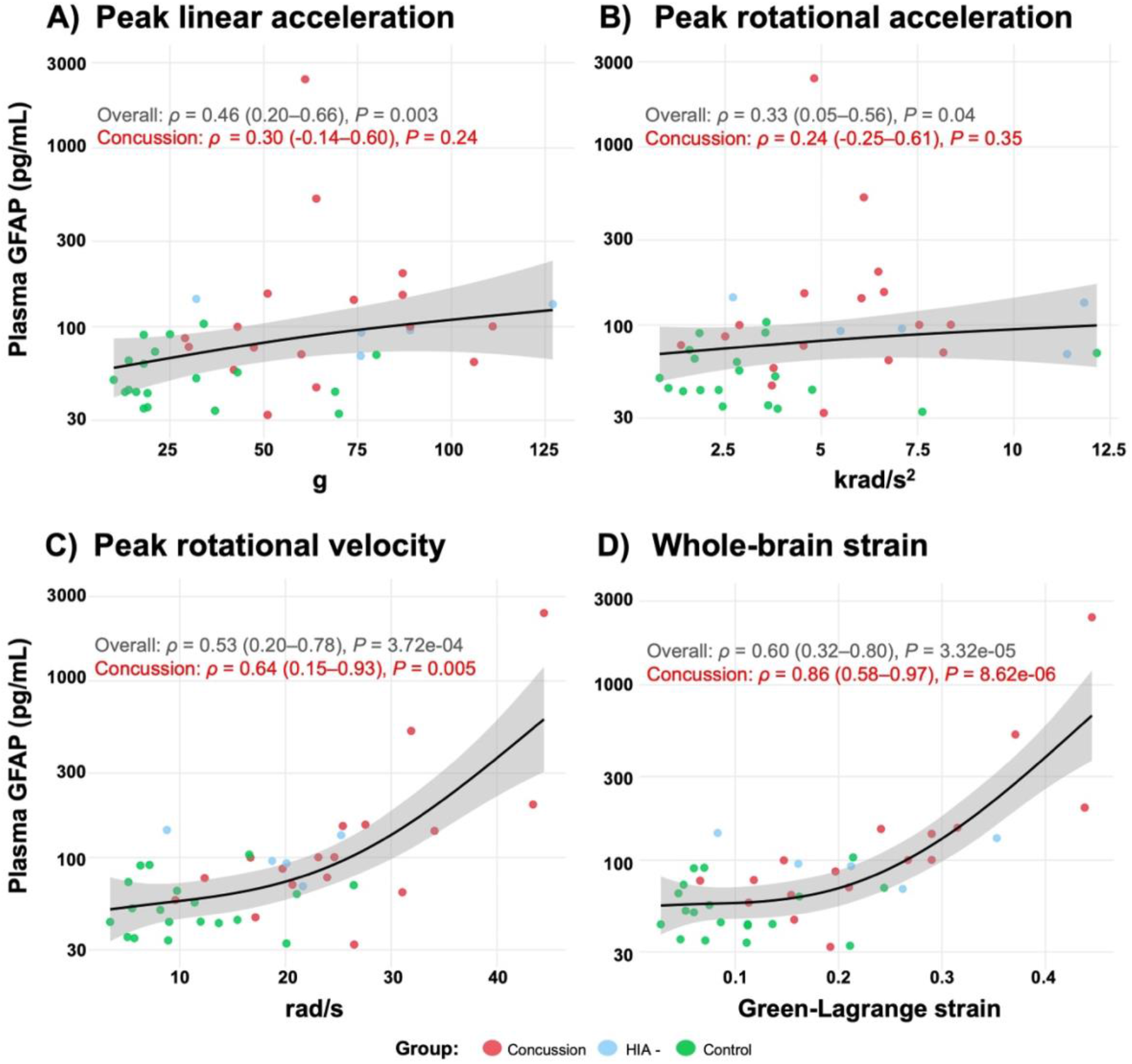
Associations between head impact biomechanics and plasma GFAP: Scatterplots show relationships between instrumented mouthguard-derived peak linear acceleration (PLA; **A**), peak rotational acceleration (PRA; **B**), and peak rotational velocity (PRV; **C**) - and finite element-estimated maximum principal Green-Lagrange strain (strain; **D**). Regional strain measures are not shown due to similar distributions to whole-brain strain; whole-brain and brainstem strain rate analyses are presented in Supplementary Figure 4. Plasma GFAP is displayed on a log_10_ scale. Trendlines were fitted using a generalised additive model with 95% confidence intervals. Spearman correlation coefficients (ρ), 95% confidence intervals and p-values are shown for the overall cohort (black; n = 41) and the concussion subgroup (red; n = 17).

**Table 1.**
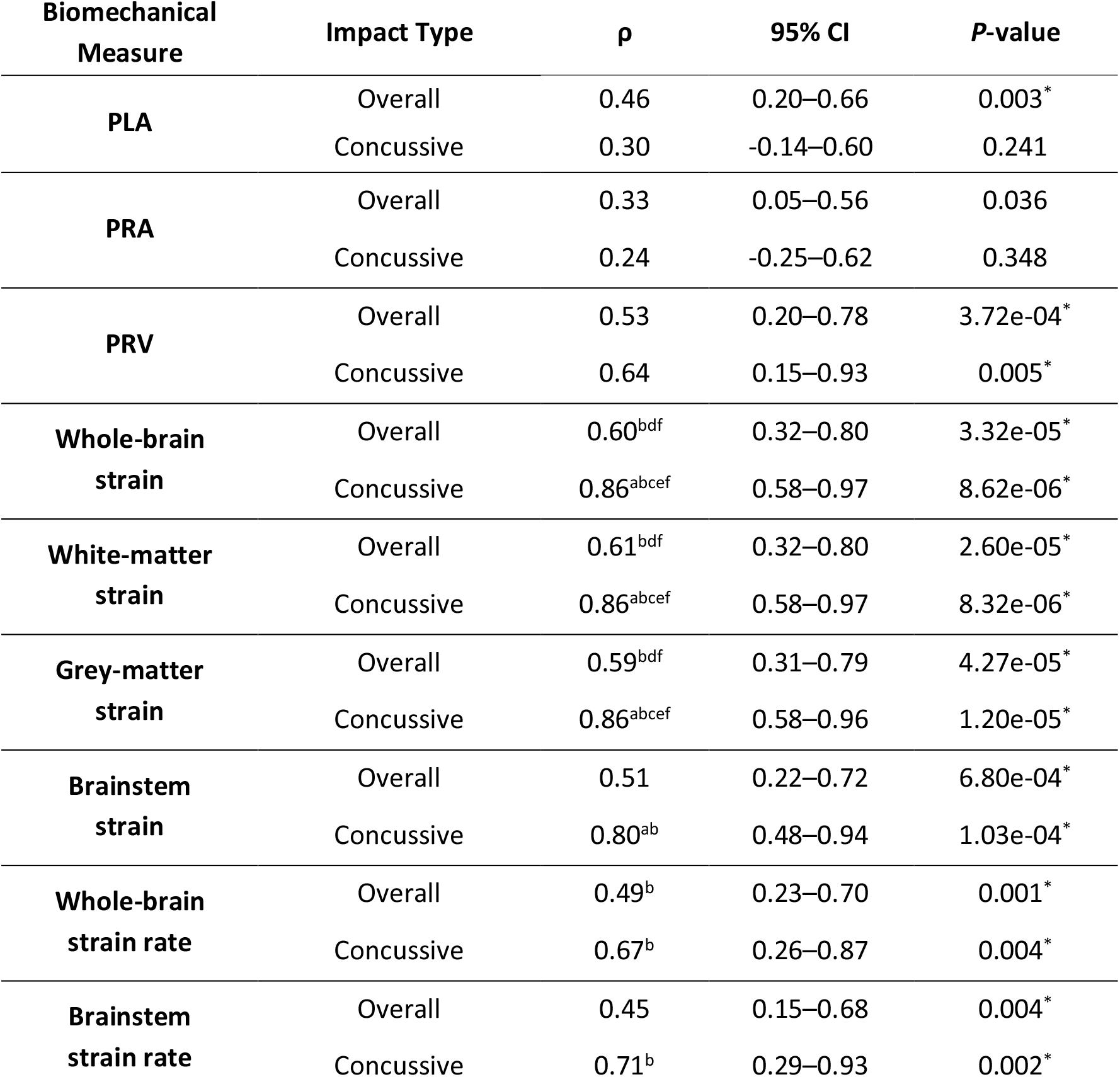
Associations between biomechanical measures and plasma GFAP. Spearman correlation coefficients (ρ) with 95% confidence intervals (CIs) and P-values are shown for the overall cohort (n = 41) and concussion (n =17) group. Biomechanical measures include peak linear acceleration (PLA), peak rotational acceleration (PRA), peak rotational velocity (PRV), and maximum principal Green-Lagrange strain (strain) in the whole-brain, white-matter, grey-matter, and brainstem; and strain rate in the whole-brain and brainstem. Superscripts indicate stronger correlations with GFAP by Steiger’s Z test: **a**, stronger than PLA; **b**, stronger than PRA; **c**, stronger than PRV; **d**,stronger than brainstem strain; **e**, stronger than whole-brain strain rate; **f**, stronger than brainstem strain rate. **\*** indicates significance after Bonferroni correction (P < 0.006).

| Biomechanical Measure | Impact Type | $\rho$ | 95% CI | P-value |
| --- | --- | --- | --- | --- |
| PLA | Overall | 0.46 | 0.20–0.66 | 0.003* |
|  | Concussive | 0.30 | -0.14–0.60 | 0.241 |
| PRA | Overall | 0.33 | 0.05–0.56 | 0.036 |
|  | Concussive | 0.24 | -0.25–0.62 | 0.348 |
| PRV | Overall | 0.53 | 0.20–0.78 | 3.72e-04* |
|  | Concussive | 0.64 | 0.15–0.93 | 0.005* |
| Whole-brain strain | Overall | 0.60<sup>bdf</sup> | 0.32–0.80 | 3.32e-05* |
|  | Concussive | 0.86<sup>abcef</sup> | 0.58–0.97 | 8.62e-06* |
| White-matter strain | Overall | 0.61<sup>bdf</sup> | 0.32–0.80 | 2.60e-05* |
|  | Concussive | 0.86<sup>abcef</sup> | 0.58–0.97 | 8.32e-06* |
| Grey-matter strain | Overall | 0.59<sup>bdf</sup> | 0.31–0.79 | 4.27e-05* |
|  | Concussive | 0.86<sup>abcef</sup> | 0.58–0.96 | 1.20e-05* |
| Brainstem strain | Overall | 0.51 | 0.22–0.72 | 6.80e-04* |
|  | Concussive | 0.80<sup>ab</sup> | 0.48–0.94 | 1.03e-04* |
| Whole-brain strain rate | Overall | 0.49<sup>b</sup> | 0.23–0.70 | 0.001* |
|  | Concussive | 0.67<sup>b</sup> | 0.26–0.87 | 0.004* |
| Brainstem strain rate | Overall | 0.45 | 0.15–0.68 | 0.004* |
|  | Concussive | 0.71<sup>b</sup> | 0.29–0.93 | 0.002* |

When stratified by group, the strongest GFAP correlations occurred in concussion cases (***Table 1***), with PRV (ρ = 0.64, 95% CI: 0.15–0.93, *P* = 0.005) and whole-brain strain (ρ = 0.86, 95% CI: 0.58–0.97, *P* < 0.001) showing moderate to strong relationships. Similar associations were seen for white-matter, grey-matter and brainstem strain, and for strain rate (***Table 1***). No associations were detected for PLA or PRA within the concussion group after multiple comparisons correction.

Steiger’s Z tests compared correlation strengths between biomechanical measures and GFAP (***Table 1, Supplementary Table 3***). Overall, GFAP correlated more strongly with strain than with PRA for whole-brain and white-matter, and grey-matter strain, and for whole-brain strain rate. Within concussion cases, whole-brain, white-matter, and grey-matter strain were consistently more strongly correlated with GFAP than PLA, PRA, and PRV; brainstem strain was more strongly correlated than PLA and PRA, but not PRV. Strain rate exceeded PRA for both whole-brain and brainstem.

### Biomechanical associations with plasma NfL

No associations were detected between kinematic or strain measures and NfL after Bonferroni correction (α = 0.006; ***Figure 5, Supplementary Table 4***). In covariate-adjusted regression models PRV (β = 0.54, 95% CI: 0.36-0.81, *P* < 0.001) and whole-brain strain (β = 0.43, 95% CI: 0.12-0.74, *P* = 0.005) were associated with NfL for impacts overall (***Supplementary Table 5***).

**Figure 5.**
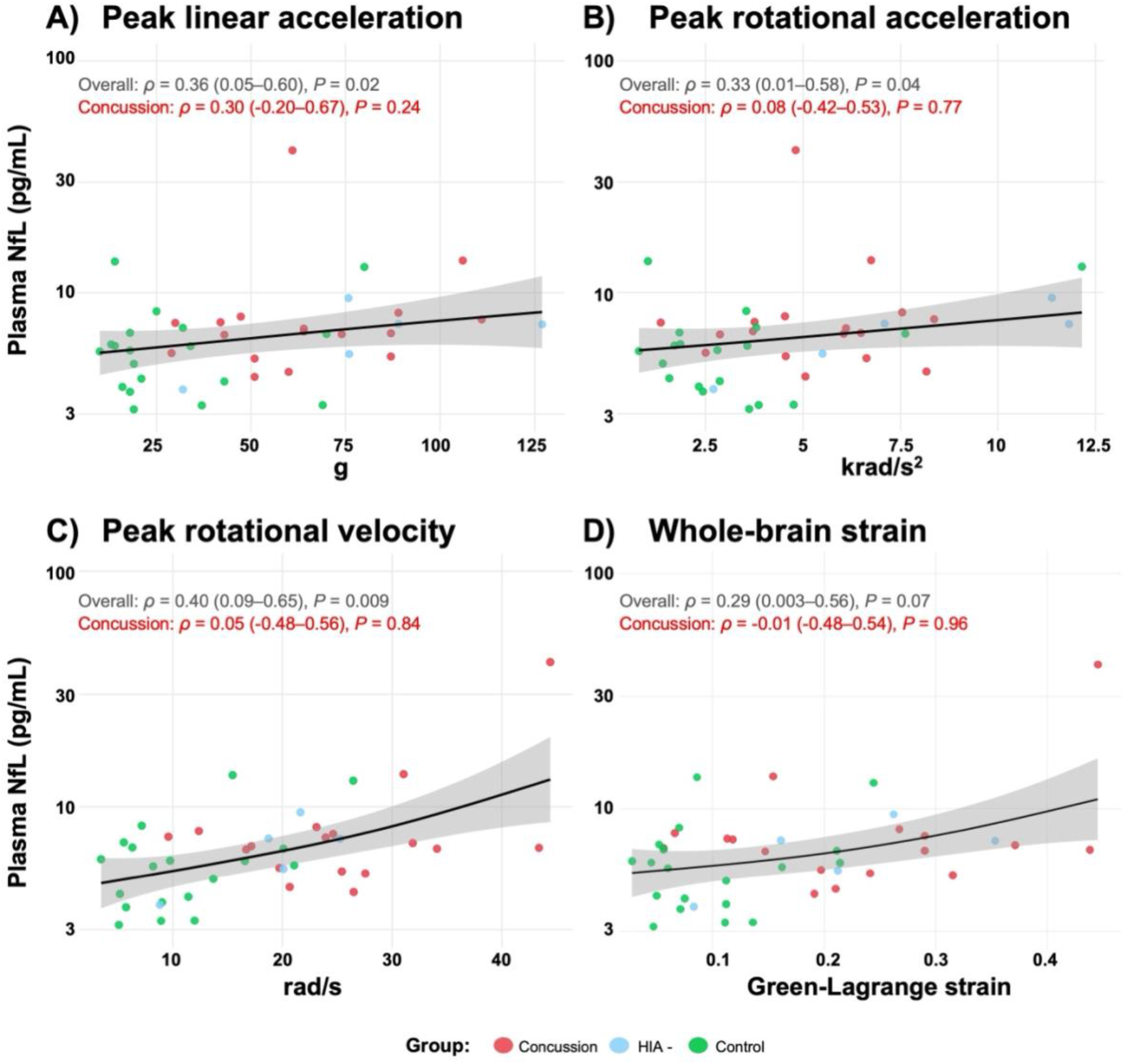
Associations between head impact biomechanics and six-day plasma NfL: Scatterplots show relationships between instrumented mouthguard-derived peak linear acceleration (PLA; **A**), peak rotational acceleration (PRA; **B**), peak rotational velocity (PRV; **C**), and finite element model-estimated maximum principal Green Lagrange strain (strain; **D**). Regional strain measures are not shown due to similar distributions to whole-brain strain; whole-brain and brainstem strain rate analyses are presented in Supplementary Figure 4. Plasma NfL is displayed on a log_10_ scale. Trendlines were fitted using a generalised additive model with 95% confidence intervals. Spearman correlation coefficients (ρ), 95% confidence intervals and p-values are shown for the overall cohort (black; n = 41) and the concussion subgroup (red; n = 17).

### Breakpoint modelling and associations with concussion symptoms

PRV showed a breakpoint at 21.0 rad/s (95% CI: 14–28) with a stronger association with GFAP above this threshold (Δslope: 0.09, 95% CI: 0.01–0.16; *P* = 0.039; ***Figure 6***). Whole-brain strain showed a breakpoint at 0.21 (95% CI: 0.15–0.27), with stronger associations beyond it (Δslope: 9.73, 95% CI: 2.60–17.01; *P* = 0.009). Similar effects were seen for white-matter (0.20, 95% CI: 0.14–0.27; Δslope: 9.80, 95% CI: 2.89–16.61; *P* = 0.008), grey-matter (0.21, 95% CI: 0.13–0.30; Δslope: 9.55, 95% CI: 2.19–16.87; *P* = 0.012) and brainstem strain (0.19, 95% CI: 0.13–0.25; Δslope: 12.20, 95% CI: 4.46–19.91; *P* = 0.034) (***Supplementary Figure 5*)**. No breakpoints were detected for PLA, PRA or strain-rate.

**Figure 6.**
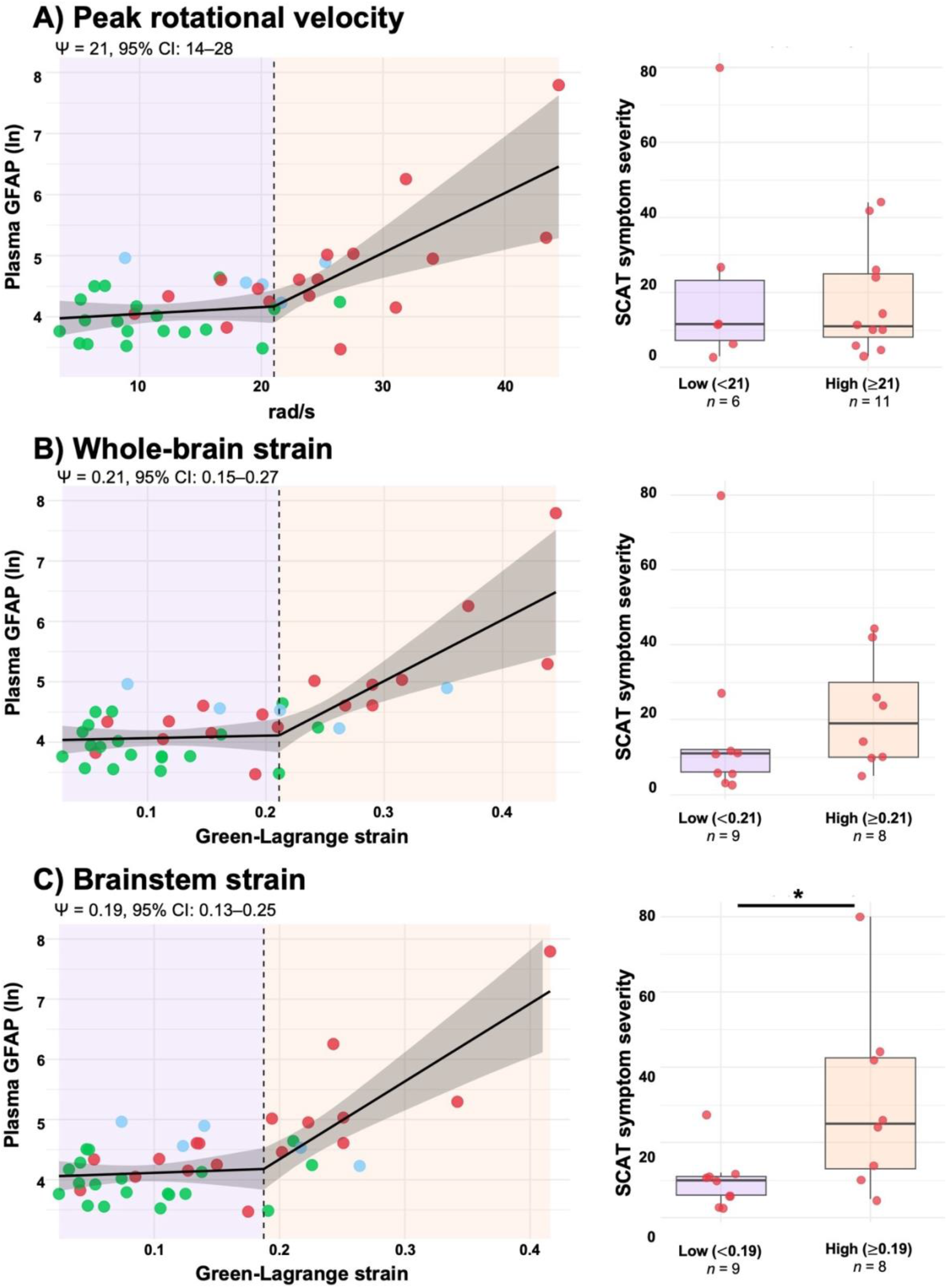
Piecewise regression analysis of biomechanics, plasma GFAP, and symptom severity: Scatterplots show relationships between peak rotational velocity(**A**), whole-brain maximum principal Green-Lagrange strain (strain; **B**), and brainstem strain (**C**) with log-transformed glial fibrillary acidic protein (GFAP). White and grey-matter strain are not shown due to similar distributions to whole-brain strain (Supplementary Figure 5). In the overall cohort, breakpoints (ψ) and 95% confidence intervals (CIs) were estimated using piecewise regression models adjusted for age, body mass index and time since injury/match. Wild bootstrapping was used to test for significant changes in slope above and below the breakpoint (P < 0.05). Points represent individual impacts from concussion (red), medically cleared head injury assessment (HIA-; blue) and control (green) groups. For measures with a significant breakpoint, differences in Sport Concussion Assessment Tool (SCAT) symptom severity within the concussion group are also shown *(P < 0.05).

To examine whether these breakpoints related to clinical outcomes in concussed players, impacts were dichotomised as above or below each breakpoint and 24 hour SCAT symptom severity scores were compared (***Figure 6***). No differences were observed for PRV (below: 11.5, IQR: 7.3–23.3; above: 11.0, 8–25; *P* = 0.920) or for whole-brain (below: 11.0, 6–12 vs above: 19.0, 10–30; *P* = 0.311), white-matter (below: 11.0, 5.3–15.8 vs above: 14.0, 10–26; *P* = 0.563) or grey-matter strain (below: 11.0, 6–12 vs above: 19.0, 10–30; *P* = 0.311). In contrast, the brainstem strain breakpoint was discriminative, with higher symptom scores above the threshold (25.0, 13–42.5) compared to those below (10.0, 6–11; *P* = 0.043).

### Cumulative impacts

Cumulative kinematic and strain measures showed no association with plasma biomarkers (***Supplementary Figure 6, Supplementary Table 6***).

## Discussion

This study aimed to advance understanding of the relationship between head-impact exposure and brain injury risk in collision sport by integrating iMG head kinematics and brain strain estimated by Imperial College brain FE model, using blood-based biomarkers as an *in vivo* ground truth for brain injury. We found that brain strain estimates showed significantly greater predictive value for plasma GFAP than peak kinematic metrics, with this relationship particularly strong within concussion cases. Piecewise regression analyses provided evidence of a biomechanical tipping point marked by a non-linear relationship with an identifiable breakpoint in brain strain above which the association with GFAP strengthened. Among concussion cases, there was evidence that impacts exceeding this strain breakpoint were associated with greater symptom burden. Taken together, these findings support the development of GFAP-anchored, FE-derived brain strain thresholds for future injury-risk models in sport.

By anchoring biomechanical measures to biomarkers as an *in vivo* ground truth, this work addresses a key limitation of prior iMG and FE validation studies that relied largely on binary clinical diagnoses^13,14^. The superior correlation of Imperial FE brain model-derived strain with plasma GFAP supports the theoretical advantage of strain-based measures, which account for the complex interplay of impact magnitude, direction, duration, and brain anatomy. Consistent with our hypothesis, these findings reinforce the utility of FE-derived strain as a more biologically relevant predictor of brain tissue damage than basic kinematics alone.

Relative to the biomechanical associations observed for plasma GFAP, corresponding associations with NfL were weak or absent. Several factors may explain this discrepancy. First, consistent with prior work, fewer concussion cases showed NfL elevations than GFAP elevations^6,27,28^, suggesting that astroglial injury/reactivity does not necessarily coincide with axonal injury. Second, NfL typically peaks around one week post-concussion and may continue to rise for several weeks in some individuals^6,27,28^, so our six-day sampling may have missed peak levels. Third, the FE model does not incorporate fibre orientation and therefore cannot capture directional vulnerability of axons to mechanical loading. Fourth, given NfL’s longer circulating half-life than GFAP, head impacts in preceding matches may have influenced concentrations. Finally, as NfL is influenced by age and BMI^36^, it is noteworthy that after adjusting for these factors in multivariate regression, PRV and strain remained significantly associated with NfL in the overall sample - the same metrics most strongly associated with GFAP. This suggests these measures may also capture aspects of axonal injury risk, potentially at higher biomechanical thresholds given the lower prevalence of NfL elevations. Larger cohorts, additional sampling timepoints, and fibre-informed modelling will be needed to better define these relationships and thresholds

While our overall dataset provided the strongest evidence that brain strain relates more closely to GFAP than kinematic metrics, the three illustrative case studies further highlight the limitations of relying on peak acceleration measures to predict brain injury. The two non-concussive cases exhibited higher peak linear and rotational accelerations than the concussive case, yet the concussive impact produced much higher FE-derived strain. The longer impact duration and higher PRV in the concussive case, together with potential differences in loading direction and site^37^, likely contributed to the higher strain and accompanying biomarker elevations.

A further advantage of FE modelling is the ability to resolve spatial patterns of brain deformation. When comparing non-concussive and concussive impacts overall, strain clusters were widely distributed, whereas strain rate clusters predominated in midline and periventricular white matter, including the corpus callosum. The broad strain distribution observed likely reflects heterogeneity in loading patterns across concussion cases. As such, FE-derived spatial information is likely to be most informative at the individual-impact level, or for quantifying cumulative regional strain exposure, or for examining how deformation in specific regions relates to clinical features. Related to this, consistent with prior work linking brainstem strain to loss of consciousness^19^, the aforementioned concussion case, which featured a prolonged loss of consciousness, showed the highest brainstem strain in the study.

Within concussion cases, brainstem strain exceeding the strain-GFAP tipping point was associated with greater symptom burden. This is biologically plausible, as the brainstem is vulnerable to biomechanical loading, and disruption of these pathways may contribute to concussion symptoms^38,39^. Similar patterns were observed for strain breakpoints in the whole-brain, white-matter and grey-matter, but not for PRV. Larger cohorts with more high-strain impacts are required to refine threshold estimates and to assess associations with symptoms.

Despite the association between exceeding the GFAP-anchored strain tipping point and greater symptom burden, GFAP should not be equated with the clinical features of concussion. Although strain-GFAP associations were strongest within concussive impacts, high-strain events with GFAP elevations were also observed in some HIA- and control cases, while some concussive cases showed low strain and GFAP levels. These patterns align with our prior studies, demonstrating good group discrimination for GFAP with partial overlap, including absent rises in some concussed athletes and increases following some non-concussive impacts^27,29,40^.

Mechanistically, plasma GFAP - a highly brain-specific astroglial protein - reflects a mixture of astrocytic injury and reactivity; with our 24 hour sampling around the GFAP peak^41^ and human^42,43^ and experimental traumatic brain injury^44–47^ data placing peak astrocyte reactivity several days later, the elevations observed here likely reflect primarily acute astrocytic injury. Astrocytic end-feet are integral to the blood-brain barrier, therefore transient GFAP elevations may reflect strain-induced damage to end-feet and short-lived barrier perturbation^48^. While such processes may contribute to symptom burden, the highly dynamic nature of concussion signs and symptoms, together with a lack of GFAP change in some concussion cases, suggests a largely physiological (e.g. neurometabolic) rather than structural basis for symptom expression. Moreover, there is considerable evidence that individual factors contribute substantially to symptom burden^24^. As such, while predicting concussion remains valuable, our working hypothesis is that models anchored to objective, brain-specific biomarkers quantified on a continuous scale such as GFAP will provide greater insight into how impact biomechanics couple to sport-related brain injury.

This study offers insights relevant to establishing best practices for using iMG data and other head impact sensors. Our findings support processing iMG-derived head kinematics through this FE brain model to generate biologically meaningful brain strain estimates. In settings where real-time output is not required, such as helmet testing, modelling may provide an optimal approach to assess brain injury risk. In contexts requiring rapid or real-time estimates, including sideline screening, machine learning models trained on larger datasets could approximate brain model-calibrated strain outputs to support timely decision-making^49^. In addition, we found that PRV, a simple metric to derive from sensor data, may offer greater predictive value than traditional acceleration measures. However, despite its practical advantages and strong association with GFAP, PRV remains a one-dimensional surrogate that does not capture the complexity of brain deformation.

Several limitations should be considered. First, we cannot exclude confounding effects of multiple impacts on biomarker levels; however, the absence of associations between cumulative exposure metrics and biomarkers supports focusing on single high-risk impacts in this setting. Second, the sample size limits power to detect more subtle associations and precludes robust subgroup analyses, particularly in non-concussive cases where the prevalence and extent of biomarker elevations are lower than in the concussion group. Third, replication in larger and more diverse cohorts is needed to confirm generalisability and enable sex-and age-specific analyses. Although the present sample provided sufficient power to detect a breakpoint, estimate precision was constrained, particularly at the upper end of the exposure where fewer high-strain impacts were observed. Future studies incorporating larger cohorts and more high-exposure events will be important to refine these estimates and better characterise biomarker trajectories around this threshold. Finally, the FE brain model used here is based on a single anatomical model and does not account for individual variability.

In conclusion, FE-modelled brain strain derived from iMGs showed stronger associations with plasma biomarkers of brain injury than traditional kinematic metrics, supporting its biological relevance to brain tissue injury. Evidence of a breakpoint in the biomechanical-biomarker relationship, together with stronger associations in concussive impacts, supports the concept of a biomechanical tipping point above which cellular injury is more likely. Collectively, these findings highlight the value of integrating brain strain into future head impact risk prediction and brain injury screening frameworks in sport.

## Statements and declarations

## Acknowledgments

We thank the management, medical staff, and players of the Victorian Amateur Football Association for their support of this study.

## Clinical trial number

Not applicable.

## Author contributions

Conceptualization: SJM, MG, JWH

Methodology: JWH, SJM, DJS, MG, KAZ

Investigation: JWH, EYKC, MG, SJM, LJE, WTO, BX, SSHR, SEB, JE, WJQZ, GS, TDP

Visualization: JWH, EYKC Supervision: SJM, SRS, MG

Writing - original draft: JWH, SJM, EYKC, MG

Writing - review & editing: All authors

