## Supplementary Materials for "Integrating mouthguard kinematics, finite element brain strain, and plasma biomarkers to explore brain injury thresholds in collision sport"

**Supplementary material:**

**Supplementary Table 1: Associations between biomechanical measures and plasma GFAP using different control impact selection criteria:**

| Control Impact Selected Based on Highest: | Biomechanical variable | ρ | 95% CI | *P*-value |
|---|---|---|---|---|
| PRA* | PLA | 0.46 | 0.20–0.66 | 0.003 |
| PRA* | PRA | 0.33 | 0.05–0.56 | 0.036 |
| PRA* | PRV | 0.53 | 0.20–0.78 | 3.72e-04 |
| PRA* | Whole-brain strain | 0.60 | 0.32–0.80 | 3.32e-05 |
| PLA | PLA | 0.40 | 0.09–0.58 | 0.011 |
| PLA | PRA | 0.34 | 0.05–0.57 | 0.027 |
| PLA | PRV | 0.46 | 0.07–0.72 | 0.003 |
| PLA | Whole-brain strain | 0.56 | 0.27–0.76 | 1.44e-04 |
| PRV | PLA | 0.60 | 0.39–0.77 | 2.97e-05 |
| PRV | PRA | 0.53 | 0.30–0.70 | 4.26e-04 |
| PRV | PRV | 0.34 | -0.02–0.66 | 0.032 |
| PRV | Whole-brain strain | 0.64 | 0.36–0.82 | 6.16e-06 |
| Whole-brain strain | PLA | 0.43 | 0.18–0.64 | 0.005 |
| Whole-brain strain | PRA | 0.35 | 0.06–0.57 | 0.027 |
| Whole-brain strain | PRV | 0.43 | 0.05–0.71 | 0.005 |
| Whole-brain strain | Whole-brain strain | 0.57 | 0.29–0.77 | 1.16e-04 |

***Supplementary Table 1: Correlation between plasma glial fibrillary acidic protein (GFAP) and biomechanical variables under different control-impact selection criteria.*** *Spearman correlation coefficients (ρ) with 95% confidence intervals (CIs) between plasma GFAP concentrations and biomechanical variables across alternative control-impact selection methods. * denotes selection method that was used in the primary analysis.*

**Supplementary Figure 1: Participant recruitment consort diagram**

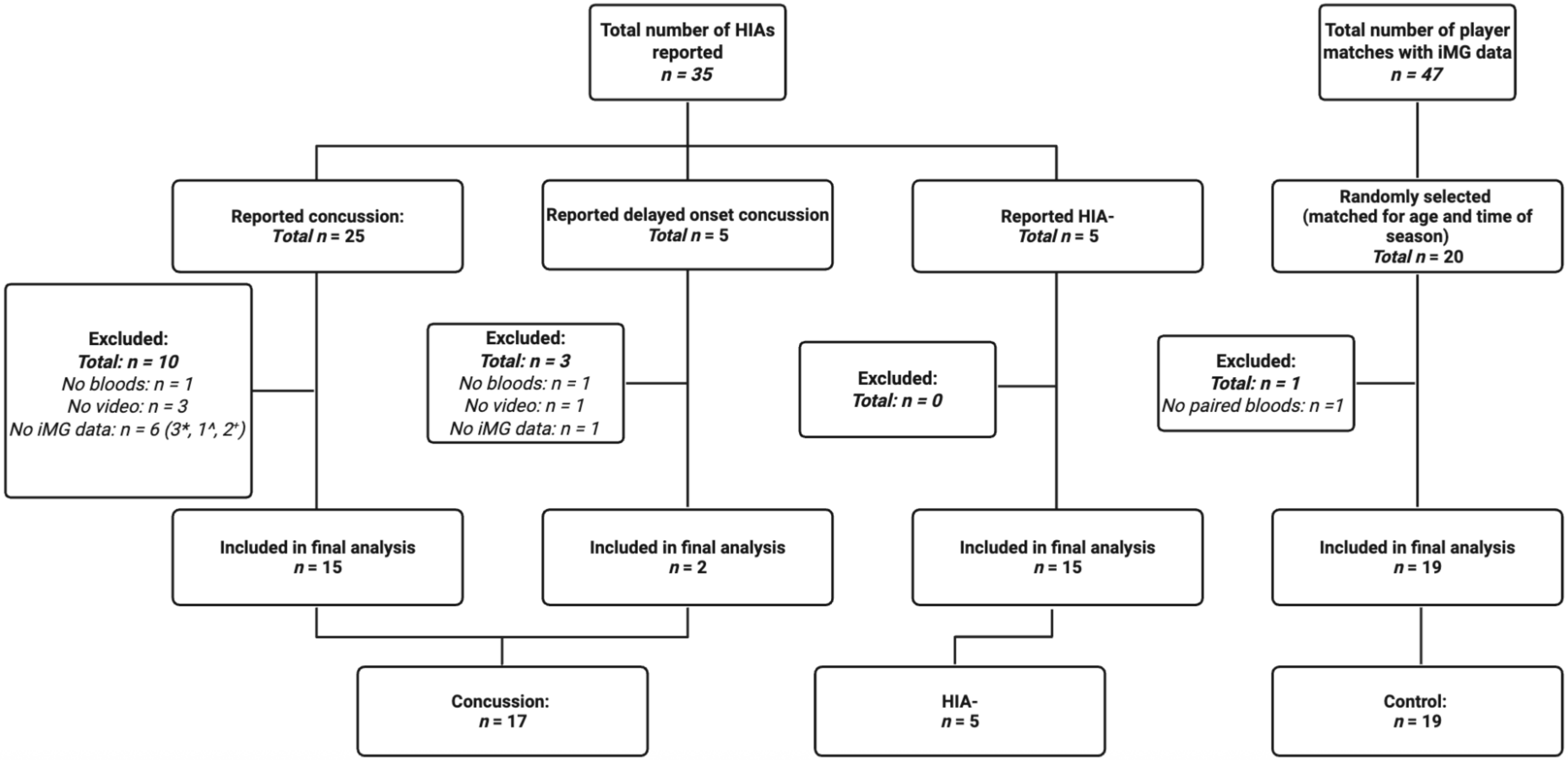


***Supplementary Figure 1: Participant recruitment flowchart.*** *Of 35 head injury assessments (HIA), 25 were concussions, 5 delayed-onset concussion, and 5 cleared assessments (HIA-). Exclusions were due to missing blood, video, or instrumented mouthguard (iMG) data (*iMGs not charged; ^no recording session created; +no data in portal). Twenty matched controls were selected; one was excluded for missing bloods. Final analyses included 41 impacts overall; 17 concussion, 5 HIA-, and 19 controls.*

**Supplementary Table 2: Demographics**

| Demographics | Concussion | HIA- | Control | *P*-value |
|---|---|---|---|---|
| Age | 23.45 (22.54– 25.21) | 25.46 (22.73–27.04) | 25.34 (24.58–27.37) | 0.16 |
| BMI | 23.89 (23.32–25.22) | 24.76 (23.27–25.17) | 24.02 (23.03–24.98) | 0.99 |
| Time since match/injury | 20.67 (19.00–22.17) | 20.82 (19.00–23.50) | 18.60 (18.00–20.13) | 0.04 |
| Number of previous concussions | 3.00 (2.00–5.00) | 4.00 (4.00–10.00) | 5.00 (3.00–7.00) | 0.53 |
| Number of impacts | 2.00 (1.00– 3.00) | 3.00 (2.00 – 4.00) | 1.00 (0.00– 4.00) | 0.52 |

***Supplementary Table 2. Participant demographics and exposure characteristics:*** *Data are median (interquartile range). P-values are from Kruskal–Wallis tests across concussion, head-injury-assessment-negative (HIA-), and control groups. Bonferroni-adjusted pairwise Mann–Whitney U comparisons were performed where appropriate (all P>0.05). BMI=body mass index. Age is reported in years; time since match/injury shown in hours.*

**Supplementary Figure 2: Distribution of biomechanical measures and plasma biomarkers**

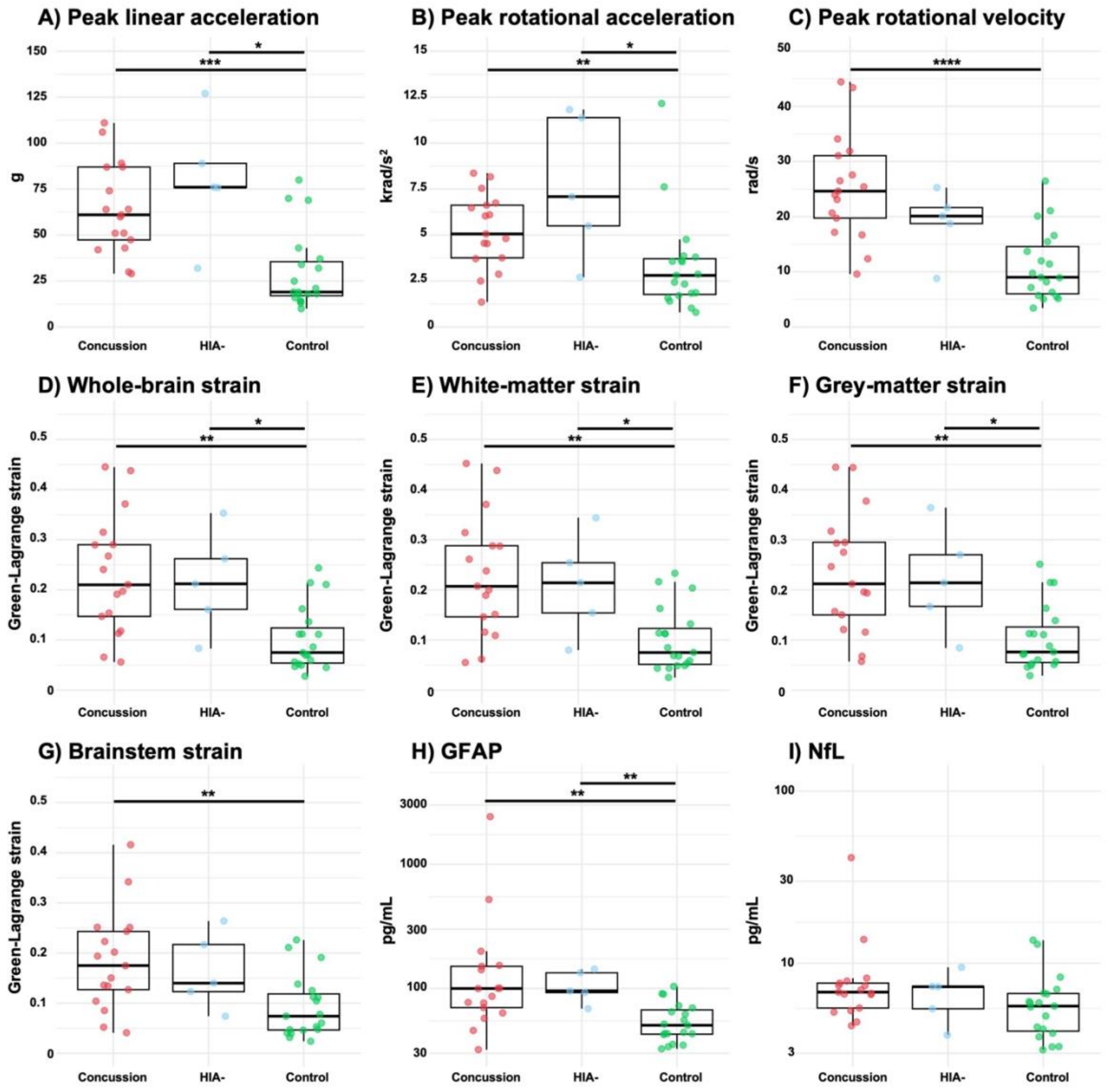


***Supplementary Figure 2: Distribution of biomechanical measures and plasma biomarkers*** *Box plots display medians and interquartile ranges; whiskers extend 1.5 x interquartile range. Dots represent individual impacts resulting in concussion (red), head-injury-assessment-negative (HIA-; blue) or the highest PRA impact from a randomly selected control match (control; green). Biomechanical measures presented from left to right include peak linear acceleration (PLA;* ***A****), peak rotational acceleration (PRA;* ***B****), peak rotational velocity (PRV;* ***C****), and 90th percentile maximum principal Green-Lagrange strain (strain), in the whole-brain (****D****), white-matter (****E****), grey-matter (****F****) and brainstem (****G****). Plasma glial fibrillary acidic protein (GFAP;* ***H****) and neurofilament light (NfL;* ***I****) levels are displayed on a $log_{10}$ scale. * $P < 0.05$, ** $P < 0.01$, *** $P < 0.001$, **** $P < 0.0001$.*

**Supplementary Figure 3: Voxel-wise group differences in brain strain and strain rate**

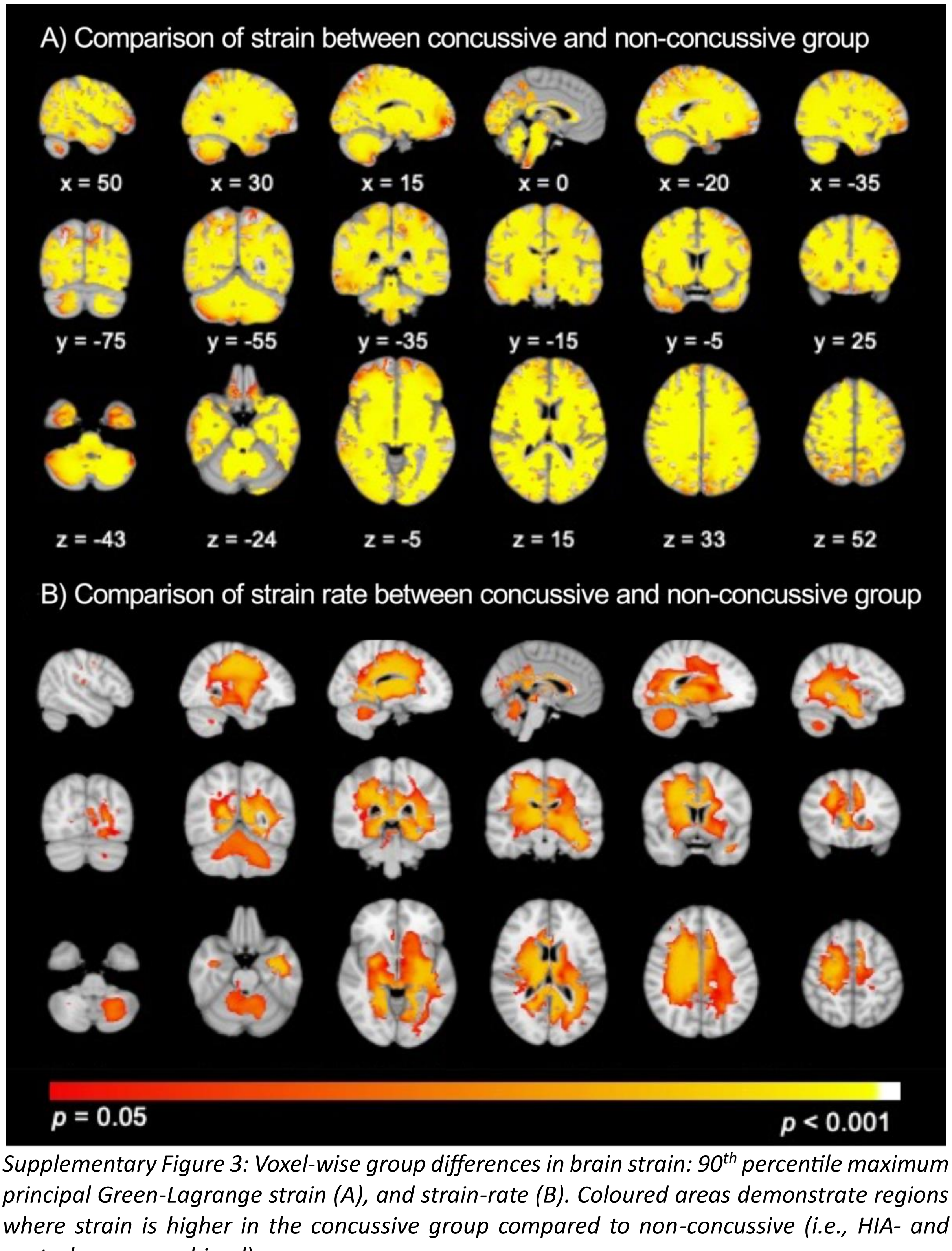


*Supplementary Figure 3: Voxel-wise group differences in brain strain: 90th percentile maximum principal Green-Lagrange strain (A), and strain-rate (B). Coloured areas demonstrate regions where strain is higher in the concussive group compared to non-concussive (i.e., HIA- and control group combined) group.*

**Supplementary Figure 4: Relationship between strain rate and plasma biomarkers**

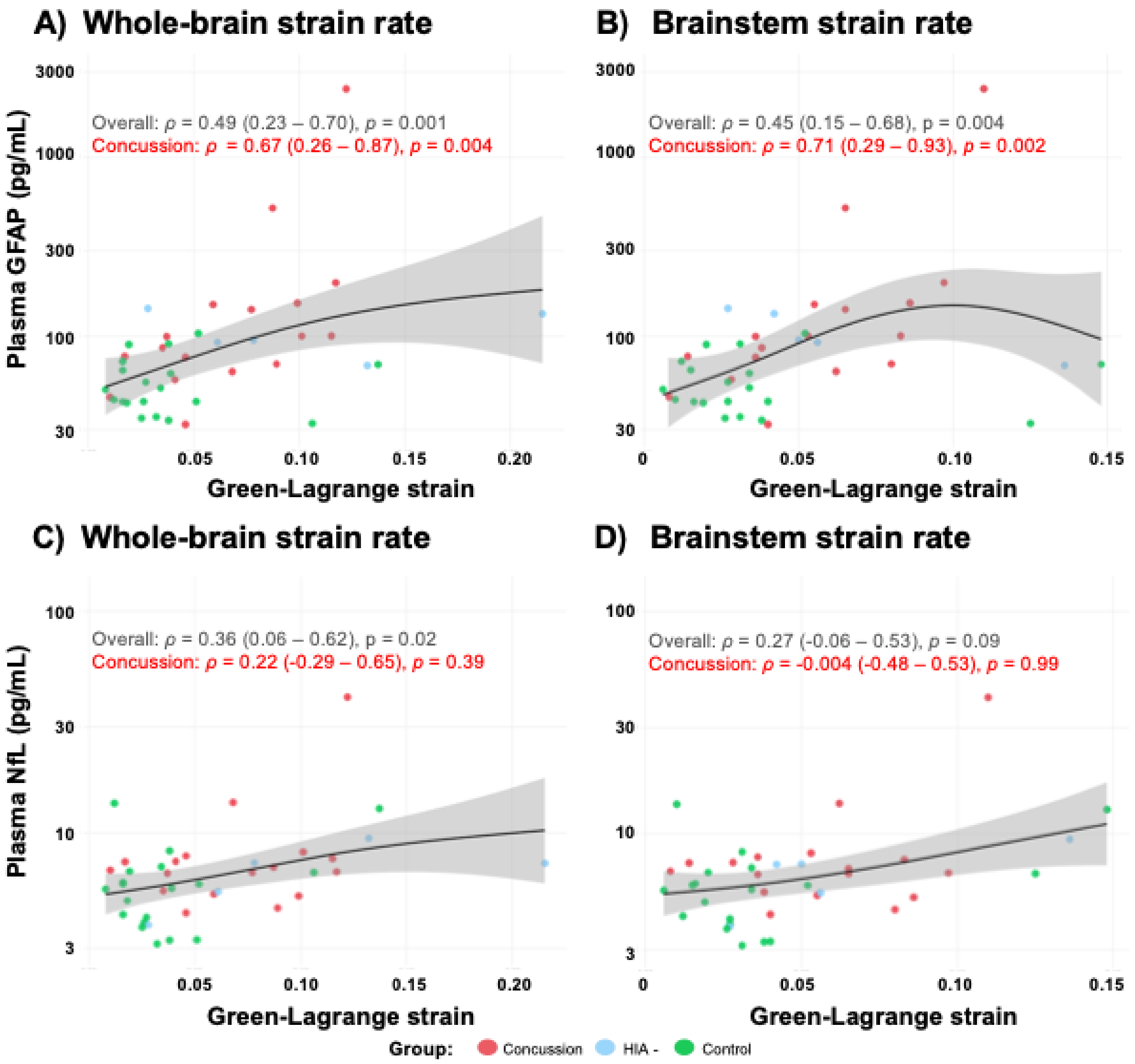


***Supplementary Figure 4: Associations between strain rate and plasma biomarker levels:*** *Scatterplots show relationships between 90th percentile maximum principal Green-Lagrange strain rate in the whole-brain (A, C) and brainstem (B, D) with plasma glial fibrillary acidic protein (GFAP) and neurofilament light (NfL) levels. Raw biomarkers levels are displayed on a $log_{10}$ scale. Trendlines were fitted using a generalised additive model for overall impacts, with 95% confidence intervals. Spearman correlation coefficients (ρ), 95% confidence intervals and p-values are shown for the overall cohort (black; n = 41) and the concussion subgroup (red; n = 17).*

**Supplementary Table 3: Comparison of GFAP correlations using Steiger Z test**

| Group | Measure 1 | Measure 2 | *Z* | *P*-value |
|---|---|---|---|---|
| **Overall** | Whole-brain strain rate | PRA | 2.96 | 0.003 |
| | White-matter strain | PRA | 2.64 | 0.008 |
| | Whole-brain strain | PRA | 2.60 | 0.009 |
| | Grey-matter strain | PRA | 2.58 | 0.010 |
| | White-matter strain | Brainstem strain | 2.46 | 0.014 |
| | Whole-brain strain | Brainstem strain | 2.32 | 0.021 |
| | White-matter strain | Brainstem strain rate | 2.11 | 0.035 |
| | Grey-matter strain | Brainstem strain | 2.08 | 0.038 |
| | Whole-brain strain | Brainstem strain rate | 2.05 | 0.040 |
| | Grey-matter strain | Brainstem strain rate | 1.99 | 0.047 |
| **Concussion** | Whole-brain strain | PRA | 3.68 | 0.0002 |
| | Grey-matter strain | PRA | 3.66 | 0.0002 |
| | White-matter strain | PRA | 3.65 | 0.0003 |
| | Whole-brain strain | PLA | 3.10 | 0.002 |
| | White-matter strain | PLA | 3.09 | 0.002 |
| | Grey-matter strain | PLA | 3.06 | 0.002 |
| | Whole-brain strain rate | PRA | 2.97 | 0.003 |
| | Brainstem strain rate | PRA | 2.83 | 0.005 |
| | Brainstem strain | PRA | 2.67 | 0.008 |
| | Whole-brain strain | Brainstem strain rate | 2.41 | 0.016 |
| | Whole-brain strain | Whole-brain strain rate | 2.38 | 0.017 |
| | White-matter strain | Brainstem strain rate | 2.38 | 0.017 |
| | White-matter strain | PRV | 2.35 | 0.019 |
| | White-matter strain | Whole-brain strain rate | 2.35 | 0.019 |
| | Whole-brain strain | PRV | 2.30 | 0.021 |
| | Grey-matter strain | Whole-brain strain rate | 2.30 | 0.022 |
| | Grey-matter strain | Brainstem strain rate | 2.28 | 0.023 |
| | Brainstem strain | PLA | 2.27 | 0.023 |
| | Grey-matter strain | PRV | 2.15 | 0.031 |

***Supplementary Table 3: Comparison of GFAP correlations using Steiger's Z test.*** *Biomechanical measures include peak linear acceleration (PLA), peak rotational acceleration (PRA), peak rotational velocity (PRV) and maximum principal Green-Lagrange strain (strain) in the whole-brain, white-matter, grey-matter, and brainstem; and strain rate in the whole-brain and brainstem.*

**Supplementary Table 4: NfL Spearman correlation table**

| Biomechanical Measure | Impact Type | ρ | 95% CI | *P*-value |
|---|---|---|---|---|
| **PLA** | Overall | 0.36 | 0.05–0.60 | 0.02 |
| | Concussive | 0.30 | -0.20–0.67 | 0.24 |
| **PRA** | Overall | 0.33 | 0.01–0.58 | 0.04 |
| | Concussive | 0.08 | -0.42–0.53 | 0.77 |
| **PRV** | Overall | 0.40 | 0.09–0.65 | 0.009 |
| | Concussive | 0.05 | -0.48–0.56 | 0.84 |
| **Whole-brain strain** | Overall | 0.29 | 0.003–0.56 | 0.07 |
| | Concussive | -0.01 | -0.48–0.54 | 0.96 |
| **White-matter strain** | Overall | 0.28 | -0.03–0.54 | 0.08 |
| | Concussive | -0.02 | -0.48–0.52 | 0.94 |
| **Grey-matter strain** | Overall | 0.29 | -0.01–0.56 | 0.06 |
| | Concussive | -0.03 | -0.48–0.49 | 0.92 |
| **Brainstem strain** | Overall | 0.19 | -0.12–0.47 | 0.22 |
| | Concussive | -0.16 | -0.62–0.44 | 0.54 |
| **Whole-brain strain rate** | Overall | 0.36 | 0.06–0.62 | 0.02 |
| | Concussive | 0.22 | -0.29–0.65 | 0.39 |
| **Brainstem strain rate** | Overall | 0.27 | -0.06–0.53 | 0.09 |
| | Concussive | -0.004 | -0.48–0.53 | 0.99 |

***Supplementary Table 4: Associations between biomechanical measures and plasma neurofilament light (NfL) concentrations.*** *Spearman correlation coefficients (ρ) with 95% confidence intervals (CIs) and P-values are reported for the overall cohort (n = 41), concussion (n =17) group. Biomechanical measures include peak linear acceleration (PLA), peak rotational acceleration (PRA), peak rotational velocity (PRV), and 90$^{th}$ percentile maximum principal Green-Lagrange strain (strain) in the whole-brain, white-matter, grey-matter, and brainstem; Strain rate is shown for the whole-brain and brainstem.*

**Supplementary Table 5: Associations between kinematic and brain strain measures with biomarker levels**

| Plasma Biomarker | Biomechanical Measure | Impact Type | β | 95% CI | *P*-value |
|---|---|---|---|---|---|
| GFAP | PLA | Overall | 0.39 | 0.08–0.71 | 0.02 |
| | | Concussive | 0.31 | -0.41–1.03 | 0.37 |
| | PRA | Overall | 0.24 | -0.09–0.57 | 0.15 |
| | | Concussive | 0.13 | -0.44–0.70 | 0.62 |
| | PRV | Overall | 0.67 | 0.43–0.92 | 2.48e-06 |
| | | Concussive | 0.68 | 0.26–1.10 | 0.004 |
| | Whole-brain strain | Overall | 0.71 | 0.47–0.94 | 4.77e-07 |
| | | Concussive | 0.72 | 0.37–1.10 | 8.00e-04 |
| NfL | PLA | Overall | 0.30 | 0.04–0.54 | 0.06 |
| | | Concussive | 0.02 | -0.42–0.80 | 0.96 |
| | PRA | Overall | 0.24 | -0.01–0.47 | 0.14 |
| | | Concussive | -0.08 | -0.37–0.46 | 0.77 |
| | PRV | Overall | 0.54 | 0.26–0.81 | 2.42e-04 |
| | | Concussive | 0.38 | -0.35–1.06 | 0.17 |
| | Whole-brain strain | Overall | 0.43 | 0.12–0.74 | 0.005 |
| | | Concussive | 0.28 | -0.44–0.92 | 0.31 |

***Supplementary Table 5: Associations between kinematic and brain strain measures with biomarker levels.*** *Multiple linear regression models for overall (n = 41), concussive (n = 17) group were adjusted for age, body mass index, and time since injury/match. Standardised beta coefficients (β) with corresponding 95% confidence intervals (CIs) and p-values are reported.*

**Supplementary Figure 5: White and grey matter regression results**

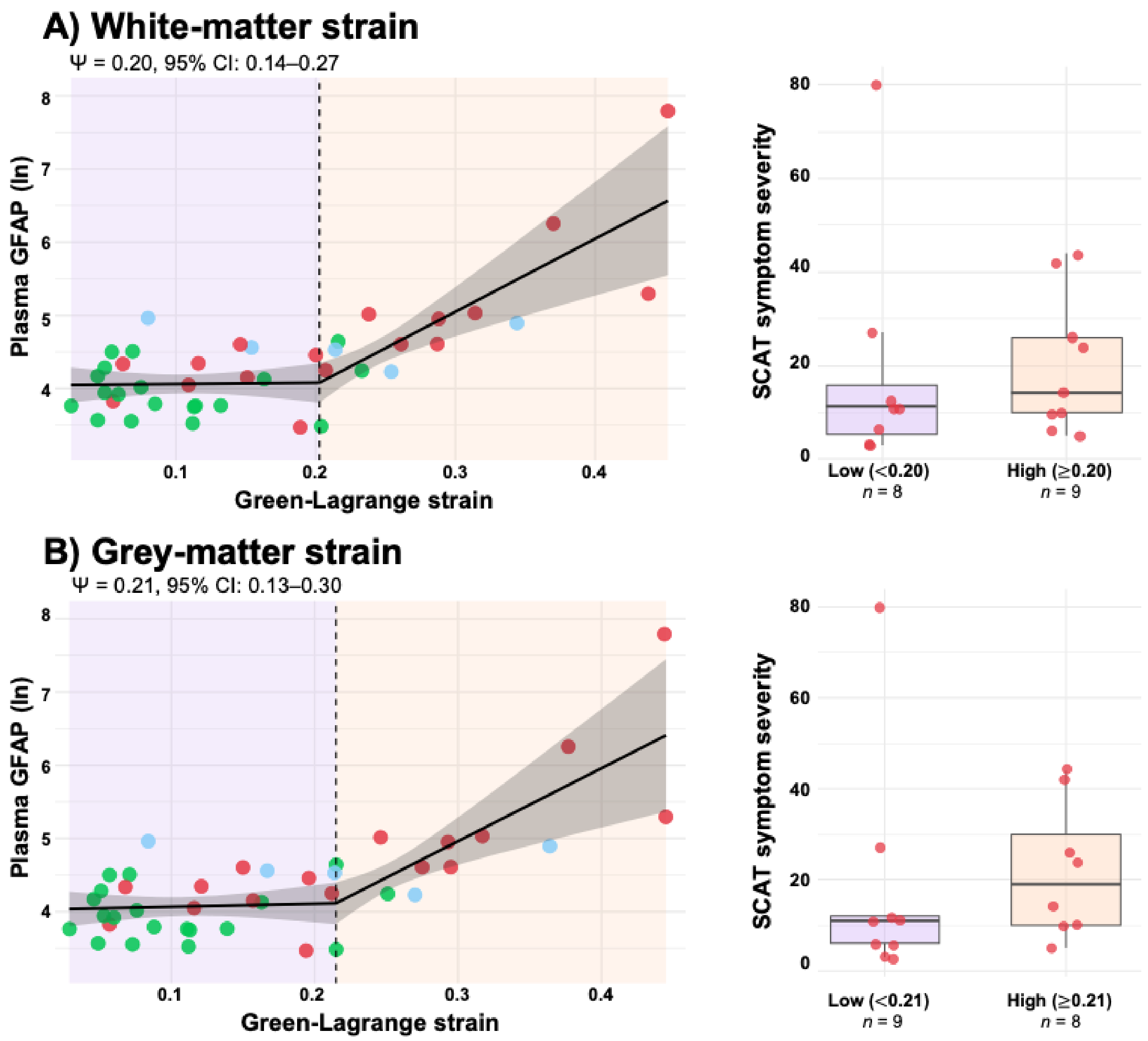


***Supplementary Figure 5: Preliminary piece-wise regression analysis between biomechanical variables, plasma biomarkers and symptom severity:*** *Scatterplots display relationships between finite element-derived 90th percentile maximum principal Green-Lagrange strain in the (**A**) white-matter and (**B**) grey-matter. In the overall cohort breakpoints (ψ) and 95% confidence intervals (CIs) were calculated using piecewise regression models adjusting for covariates age, BMI and time since injury/match. Wild bootstrapping was performed to calculate whether the change in slope was significant above and below this point ($P < 0.05$). Individual data points show single impacts from concussion (red), medically cleared head injury assessment (HIA-; blue) and control (green) groups. Differences in sport concussion assessment tool, 5th edition (SCAT) symptom severity within the concussion group is also displayed for each measure with a significant breakpoint.*

**Supplementary Figure 6: Cumulative head impact exposure biomarker associations**

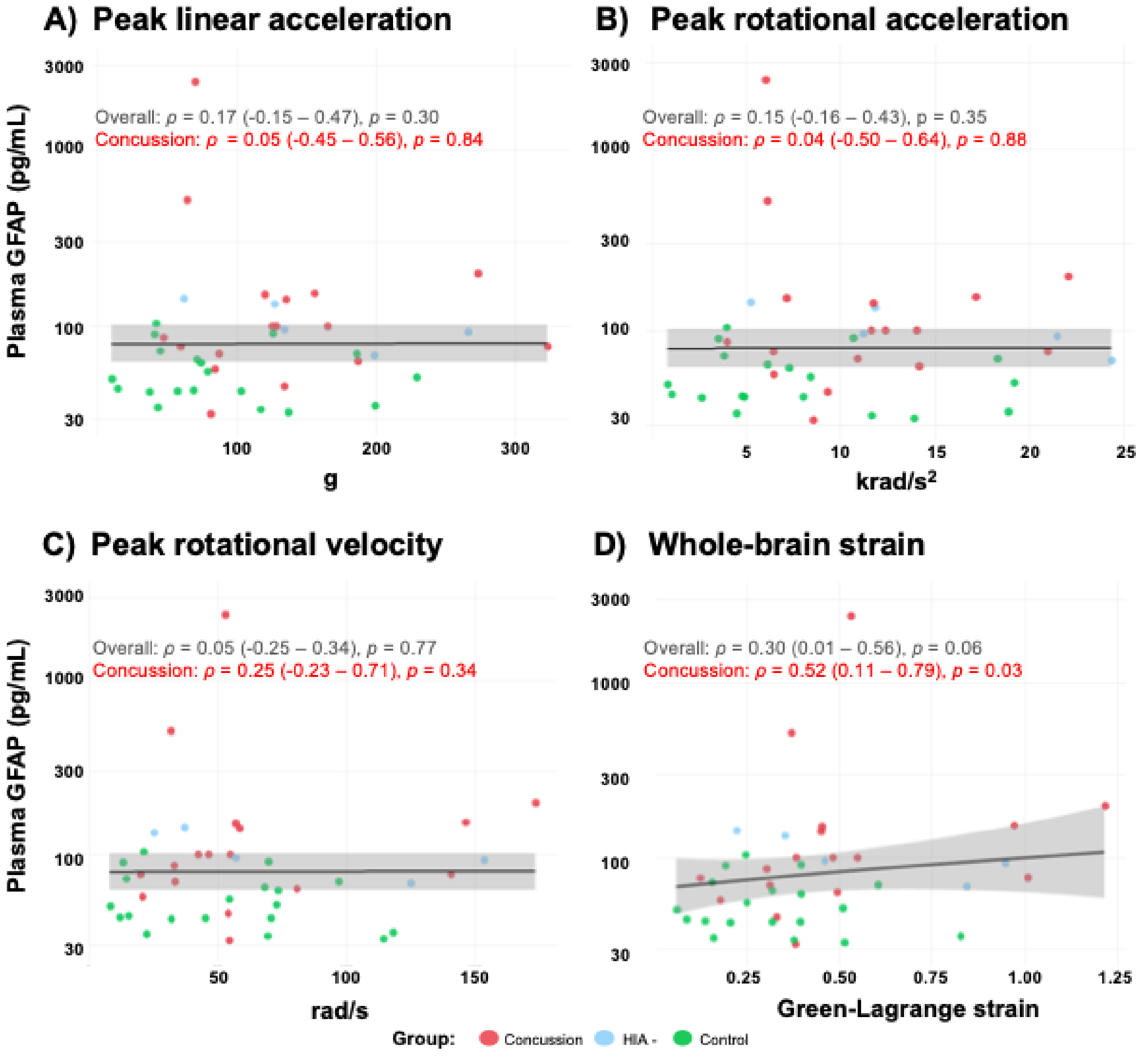


***Supplementary Figure 6. Relationship between cumulative head impact metrics and glial fibrillary acidic protein (GFAP) levels**. Scatterplots show relationships between cumulative instrumented mouthguard-recorded kinematics over a single match - (**A**) summed peak linear acceleration (PLA), (**B**) summed peak rotational acceleration (PRA), and (**C**) summed peak rotational velocity (PRV) - alongside finite element model-estimated maximum principal Green-Lagrange strain (Strain; **D**). Raw GFAP levels are displayed on a $\log_{10}$ scale. Trendlines were fitted using a generalised additive model for overall impacts, with 95% confidence intervals. Spearman correlation coefficients (ρ), 95% confidence intervals and p-values are shown for the overall cohort (black) and the concussion subgroup (red).*

**Supplementary Table 6: Correlations and associations between cumulative biomechanical measures and plasma biomarkers**

| Plasma Biomarker | Biomechanical Measure | Impact Type | ρ | 95% CI | *P*-value |
|---|---|---|---|---|---|
| **GFAP** | **PLA** | Overall | 0.17 | -0.15–0.47 | 0.30 |
| | | Concussion | 0.05 | -0.45–0.56 | 0.84 |
| | **PRA** | Overall | 0.15 | -0.16–0.43 | 0.35 |
| | | Concussion | 0.04 | -0.50–0.64 | 0.88 |
| | **PRV** | Overall | 0.07 | -0.25–0.34 | 0.77 |
| | | Concussion | 0.25 | -0.23–0.71 | 0.34 |
| | **Whole-brain strain** | Overall | 0.30 | 0.01–0.56 | 0.06 |
| | | Concussion | 0.52 | 0.11–0.79 | 0.03 |
| **NfL** | **PLA** | Overall | 0.29 | -0.04–0.57 | 0.07 |
| | | Concussion | 0.08 | -0.40–0.55 | 0.77 |
| | **PRA** | Overall | 0.27 | -0.05–0.55 | 0.09 |
| | | Concussion | -0.01 | -0.51–0.54 | 0.97 |
| | **PRV** | Overall | 0.13 | -0.20–0.42 | 0.44 |
| | | Concussion | -0.14 | -0.56–0.33 | 0.60 |
| | **Whole-brain strain** | Overall | 0.30 | -0.03–0.61 | 0.06 |
| | | Concussion | 0.16 | -0.35–0.62 | 0.55 |

***Supplementary Table 6: Associations between cumulative kinematic and brain strain measures with plasma biomarker levels.*** *Spearman correlation coefficients (ρ) with 95% confidence intervals (CIs) and p-values are reported for cumulative (summed) peak linear acceleration (PLA), peak rotational acceleration (PRA), peak rotational velocity (PRV), and whole-brain strain measures in the overall cohort (n = 41) and concussion (n = 17) group, with plasma glial fibrillary acidic protein (GFAP) and neurofilament light (NfL) levels.*
